# Validity of the Web-based, Self-directed, NeuroCognitive Performance Test in MCI

**DOI:** 10.1101/2021.10.29.21265565

**Authors:** P. Murali Doraiswamy, Terry E. Goldberg, Min Qian, Alexandra R. Linares, Adaora Nwosu, Izael Nino, Jessica D’Antonio, Julia Philips, Charlie Ndouli, Caroline Hellegers, Andrew M. Michael, Jeffrey R. Petrella, Howard Andrews, Joel Sneed, Davangere Devanand

## Abstract

Digital cognitive tests offer several potential advantages over established paper-pencil tests but have not yet been fully evaluated for the clinical evaluation of mild cognitive impairment. The NeuroCognitive Performance Test (NCPT) is a web-based, self-directed, modular battery intended for repeated assessments of multiple cognitive domains. Using a sample of 101 MCI subjects, we report in this study that the NCPT composite is significantly correlated with both a composite measure of established tests (r=0.77, p<0.0001) as well as with the ADAS-Cog (r=0.55, p<0.0001). Both test batteries had a similar factor structure that included a large “g” component with a high eigenvalue. Further, both the NCPT and established tests significantly (p< 0.01) predicted the UPSA and FAQ, measures of daily functioning. Despite limitations such as a relatively small sample, absence of control group and cross-sectional nature, these findings are consistent with the growing literature on the promise of self-directed, web-based cognitive assessments for MCI.

## INTRODUCTION

The advent of cloud-based online and smartphone cognitive tests has enhanced access to home-based and point of care memory evaluation (1-3). There has been a recent proliferation of such tests including several that have been cleared by regulatory agencies (4-13). Such tests offer many potential advantages over conventional clinician-administered neuropsychological tests such as greater consistency of administration, easy generation of alternate forms, 24/7 access on demand, better stimulus control, greater ability to test at home, greater ability to personalize and minimize floor or ceiling effects, automation of scoring, cloud storage and integration with electronic health records, and ability to scale to diverse populations and settings, such as areas with clinician shortages (12). Indeed, the computerized version of the ADAS-Cog has been shown to have better reliability than the traditional paper-pencil version (13). Further, the pandemic has also raised the need for remote contactless assessment both for clinical purposes and clinical trials in elderly at-risk patients (1). There are also potential disadvantages of computerized tests in that they can be difficult for more severely impaired individuals or those unfamiliar with computers and are prone to technical glitches and privacy breaches. These issues argue for further study in diverse clinical settings to determine the best tests for specific purposes (14).

The NeuroCognitive Performance Test (NCPT) is an online, digital platform intended for repeated assessments of multiple domains such as different types of attention, memory, executive functioning and psychomotor speed (3). It currently has 18 modules (subtests), each based on well-known neuropsychological assessments and its modular design allows researchers to develop customized batteries to address specific research outcomes. The NCPT is a self-directed test designed to be taken remotely in an unsupervised fashion through a web browser. Morrison et al (3) reported on normative data and factor structure of an 8-item NCPT in 130,140 healthy volunteers, drawn from 187 countries and across a wide educational and age range. They reported adequate test-retest reliability over 70 days (in a subset of 35,779 users) as well as good concurrent validity to standard neuropsychological tests (in a subset of 73 younger subjects). Compared to age-, gender-, and education-matched normal controls, the NCPT composite was 0.78 SD lower in subjects with self-reported mild cognitive impairment (MCI) (N=1473) and 1.17 SD lower in subjects with self-reported Alzheimer’s disease (AD) (N=105) (3). In a subsequent study of 4715 subjects, a 7-item version of the self-directed NCPT was sensitive to detecting the effects of cognitive training (15). These findings support further study of the utility of the NCPT in clinical populations.

In this study, we report our cross-sectional experience using the NCPT at baseline in a two-site, prospective clinical trial of MCI (16). A 10-item version of the NCPT was created for this study incorporating modules thought to be sensitive to the study interventions (16). The aims of this paper were to evaluate the feasibility of NCPT-10 self-administration and examine its construct against standard paper-pencil tests.

## METHODS

### Subjects and Study design

All subjects gave written informed consent and the study was approved by the respective institutional IRBs. Details of study design, inclusion exclusion criteria and study procedures have been previously reported (16). 109 MCI subjects were recruited for a prospective clinical trial of computerized brain training at two study sites (New York and Durham) stratified by MCI severity (early MCI or late MCI) and age (70 and below or 71 and above). One hundred and one subjects with completed baseline NCPT and standard neuropsychological test data were included in this analysis. A clinical diagnosis of MCI was made after neuropsychiatric and neuropsychological evaluation as described previously (16). All subjects also underwent brain MRI. Severity of MCI was assessed by the delayed recall score of WMS-III Logical Memory as described previously (16). Notable inclusion criteria included an age range of 55–95 years, subjective cognitive complaints (ie, memory or other cognitive complaints, eg, naming/language), a Wechsler Memory Scale-III (WMS-III) Logical Memory delayed recall score 0-11 (education adjusted), Mini Mental State Examination (MMSE) score ≥23 out of 30, availability of an informant and access to a home desktop or laptop computer with full access to the Internet for the study duration. Notable exclusion criteria included major neuropsychiatric illness, lack of English-speaking ability, and regular use of brain training games (16).

### Neuropsychological and Functional Assessments

At baseline, the Alzheimer’s Disease Assessment Scale-Cognition Subscale 11 (ADAS-Cog 11) was administered, followed by the following neuropsychological test battery: Digit Symbol Substitution Test (DSST) (to assess attention), WMS-III Visual Reproduction Test (to assess nonverbal learning and memory), Auditory-Verbal Learning Test (to assess verbal learning and memory), Block design, Verbal and Category Fluency, Trail Making A & B (to assess attention and executive function), and 15-item Boston Naming Test (to assess language) (16). For word learning lists, the neuropsychological testing materials provided different but parallel word lists, so as to avoid practice effects in MMSE and ADAS-Cog, but not for AVLT because we were concerned that different AVLT forms had not been established as equivalent in difficulty level (16). Following this test battery, the University of California San Diego Performance-Based Skills Assessment (UPSA-3) and the 40-item, scratch and sniff, University of Pennsylvania Smell Identification Test (UPSIT) were also administered (16-18). The UPSA-3 is a performance-based measure of cognitive and functional abilities that includes measures of simulated real-world activities; for example, planning a trip to the beach, remembering documents to bring to a medical appointment, and dialing a phone number (17). The Functional Activities Questionnaire (FAQ) was administered to the patient’s informant, either during the study visit or shortly after the visit over the phone. Testing fatigue was mitigated by allowing participants to take breaks during the testing.

### NeuroCognitive Performance Test (NCPT-10)

Following the paper-pencil testing session, subjects were then taken to a quiet clinic room so they could do the NCPT (Lumos Labs) by themselves in a self-directed manner. The study provided the on-site computer for this test. Subjects had their own study specific log-ins. Although the NCPT was designed to be an unsupervised test, in this study we had a research associate available to help troubleshoot if computer glitches arose. The cognitive domains measured by the NCPT were memory (visuospatial working memory, short-term memory), processing speed (visual search, psychomotor speed), problem solving (logical reasoning, numerical calculation), attention (selective, divided) and flexibility (response inhibition, task switching). The 10 NCPT subtests were online adaptations of widely used neuropsychological tests (12, 16, Supplemental Table 1). The NCPT took about 20-40 minutes.

**Table 1.** Baseline Characteristics of MCI Subjects*. (mean ± SD)

| **Label** | **Total** |
| --- | --- |
| N | 101 |
| NYC / Durham (% of sample) | 52 / 49 |
| Age (years) | 70.8 ± 8.9 |
| Female/Male (%) | 58.4 / 41.6 |
| White / Minority (%) | 76.2 / 23.8 |
| Education (years) | 16.7 ± 3.1 |
| EMCI / LMCI (%) | 44 / 57 |
| MMSE Total | 27.1 ± 1.7 |
| Wechsler Logical Memory Delay | 6.4 ± 3.3 |
| ADAS-Cog-11 Total | 9.5 ± 3.5 |
| UPSA Total | 80.9 ± 11.3 |
| FAQ Total | 3.4 ± 4.0 |
| UPSIT Total | 28.2 ± 7.7 |

\*Please see text for details on sample size and abbreviations

### Statistical Approaches

In this paper our goal was to examine the cross-sectional relationship, at baseline, of established tests (9 domain-specific neuropsychological tests and 2 global cognitive tests [ADAS, MMSE]) to the 10-item NCPT as well as the NCPT’s relationship to measures of daily function (the UPSA-3 and FAQ), disease severity, and odor identification. Summary statistics for demographic and various tests were calculated. Spearman correlation were constructed to examine the interrelations of the various tests. For the NCPT and the paper-pencil tests separately, we conducted a principal components analysis followed by a factor analysis (varimax rotated). For the principal components we determined the number of factors by setting eigen values>1 and by scree plots. We considered an individual test to be associated with a specific factor if its loading was greater than 0.40. Linear and logistic regressions examined the association of the factors with functional measures and disease severity. We also constructed an exploratory composite Z score for the NCPT tests weighting the 10-tests equally, as well as composite z-score for the key paper-pencil tests, and examined their association with the functional measures and disease severity. For computing the composite z-scores for both established and NCPT batteries, we reverse coded (reversing sign) ADAS-Cog, Trails-A and Trails-B, so that higher numbers for all tests reflect better performance. As such, this was an exploratory analysis and we did not adjust the p-values for multiple comparisons.

## RESULTS

Table 1 depicts the baseline characteristics of the 101 MCI subjects. Of these, 44 subjects were classified as EMCI and 57 as LMCI. Subjects recruited at Durham were on average older than those recruited in NYC but there were no differences in gender ratio or educational level. The mean ADAS, MMSE, UPSA and FAQ scores of our subjects are consistent with those reported in other MCI clinical trials.

### Correlations between NCPT and Paper-Pencil tests

Within the NCPT and traditional test batteries, there were multiple significant test intercorrelations, suggesting the potential presence of a general cognitive ability factor (or g) factor in each (data not shown). As shown in Table 2, there were multiple statistically significant correlations between the NCPT and paper-pencil tests. The correlation between the composite of all paper-pencil tests (CompZ) and the NCPT composite (NCPT-Z) was high (r=0.78, p<0.0001). The correlation for the analogous tests, Trails A (r=0.43) and Trails B (r=0.63) between the NCPT and the paper-pencil versions were significant (p<0.001). Likewise, the correlations between the NCPT word list learning memory tests and AVLT were significant (p<0.0001). The correlations between NCPT composite Z score and ADAS (r=-0.55, p<0.0001) (Figure 1) and MMSE (r=0.56, p<0.0001) were also significant.

**Table 2.**
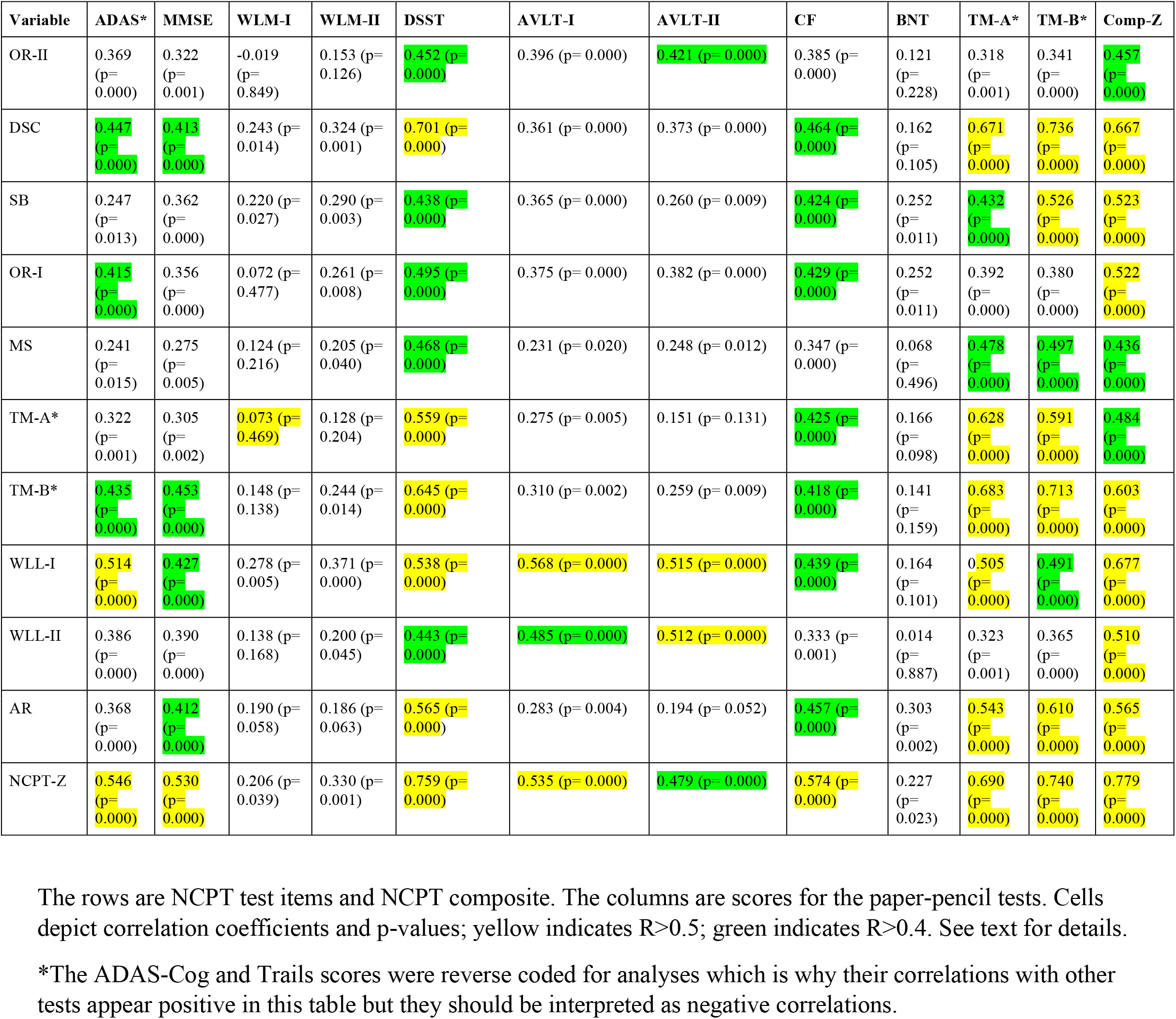
Spearman correlation (p-value) between established and NCPT measures.

**Figure 1:**
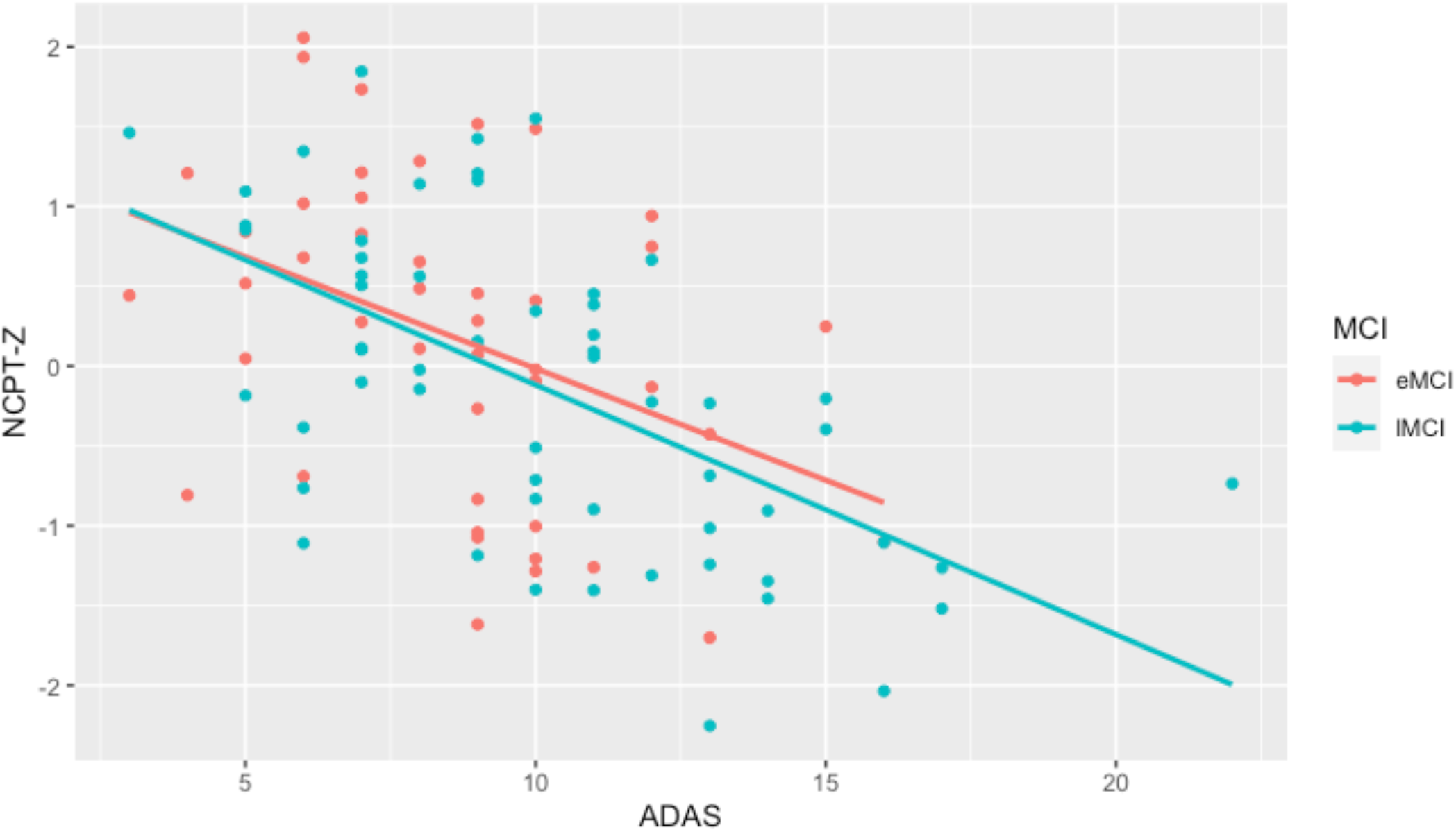
Correlation between NCPTz and ADAS-Cog in MCI. Figure1 shows correlation between NCPT-Z composite and ADAS-Cog. The red and blue regression lines show correlations for EMCI and LMCI subjects respectively. Raw ADAS-Cog scores (not reverse coded) were used for this figure. See text for details.

### Principal Components and Varimax Analyses of NCPT and Paper-Pencil tests

Three factors were derived for both the NCPT and paper-pencil measures (Supplemental Table 2). For the NCPT the first component accounted for 46% of the variance and was comprised of multiple tests from different domains (e.g., scale balance reasoning, trails speed, arithmetic). The second component was comprised of word list learning scores (verbal recall) as well as digit symbol coding and Trails. The third was comprised of the two object recognition memory scores. The three components accounted for 69% of the variance. For the paper-pencil tests, the component factor with the largest eigenvalue included tests of memory, general tests (MMSE and ADAS-Cog) and speed, which could represent “g” (Supplemental Table 2). It accounted for 44% of the variance. A second factor was comprised of high loadings from the AVLT delayed list learning and delayed logical memory. The third factor included logical memory immediate and naming, suggesting a verbal recall construct. All three components accounted for 68% of the variance, which was similar to the 69% for the NCPT.

The first factors in each battery were highly inter-correlated even after adjusting for age, gender and education (r=0.60, p<0.0001). This suggests they both assay a latent construct involving general cognitive ability (“g”). The second factors in each battery were also significantly inter-correlated (r=0.35, p<0.0004) after adjustment for age, education and gender. The third factor in each battery was not inter-correlated (p<0.33).

### Relationship of Factors to Daily Functioning and Olfactory Deficits

We examined the relationship of these factor scores to daily functioning (FAQ, UPSA), the UPSIT odor identification test known to predict AD, and severity of MCI (EMCI vs LMCI). The results below report values for regressions after adjusting for age, sex, and education. For the NCPT the UPSIT was predicted significantly by factors 2 (p<0.002) and 3 (p<0.0001), the UPSA by factors 1 (g), 2, and 3 (p<0.0001 for all), and the FAQ by factors 1 (g) (p<0.002) and 2 (p<0.0001). No NCPT factor predicted early MCI/late MCI membership. For the established measures, the UPSIT was predicted primarily by factor 1 (g) (p<0.007), the UPSA by factors 1, 2 and 3 (p<0.0001) and the FAQ by factors 1 and 2 (p<0.000). For the established measures, MCI severity was predicted by factors 2 and 3 (p<0.0001). For the two functional measures (UPSA and FAQ), the R^2^ and pseudo-R^2^ accounted for by both NCPT and paper-pencil tests were much larger than that accounted for by demographics. However, for the UPSIT, the R^2^ accounted for by NCPT and demographics was larger than that accounted for by the paper-pencil tests. Alternate models with all factors or 5-factors were also examined but not deemed superior (data not shown). A separate regression analyses (not using principal factors) confirmed that NCPT and paper-pencil composite scores both significantly predicted the UPSA and FAQ (data not shown).

## DISCUSSION

The NCPT, a self-directed, web-based, computerized test, has one of the largest normative databases of its kind derived from over 40 million users in a natural setting (19). However, in contrast to some other computerized tests, such as NTB (20), CANTAB, Cogstate or CNS Vital Signs, it has not been used as extensively in memory clinics or in MCI/AD clinical trials. In this analysis, we compared the NCPT with a paper and pencil battery in subjects with clinically diagnosed MCI.

Several key findings emerged from our study. The feasibility of completing the self-directed NCPT was high among MCI subjects. Both test batteries had a similar factor structure that included a large “g” component with a high eigenvalue. Both test batteries also showed broadly similar factor score prediction of two clinically meaningful functional outcomes. Both NCPT and established test batteries were also significantly associated with FAQ and UPSA, which are established, clinically meaningful, functional measures (17). The NCPT was also associated with the UPSIT, a test of olfactory function that has been shown to predict future cognitive decline (18). Analogous tests in the NCPT and paper-pencil batteries, such as Trails and word list learning, were highly correlated. Likewise, all ten NCPT tests as well as an NCPT z-score composite were significantly correlated with the ADAS-Cog, an instrument used widely in clinical trials. All ten NCPT items and the composite were also correlated with the MMSE, a screening tool widely used in clinical practice. These data support the concurrent validity of the NCPT.

The strengths of our study are the clinical verification of MCI diagnosis and standardized administration of a comprehensive battery of neuropsychological tests on the same day as the NCPT. However, there were also some limitations to our study. Our analysis was cross-sectional and lacked a healthy control group; hence, we could not directly assess the utility of NCPT as a diagnostic or prognostic tool. The NCPT modules in this study were selected to examine intervention effects and as such were not intended to be exactly analogous to the paper-pencil tests. This was taken into account in conducting the factor analyses and did not affect the results materially with factor 1 and factor 2 being similar for the paper-pencil battery and NCPT. We did not measure biomarkers and hence cannot determine the cause of MCI. We did not adjust for multiple comparisons in our correlation analyses and hence these results should be viewed as preliminary.

In summary, our analysis finds that the NCPT, a web-based, self-directed, computerized test, shows high concurrent validity with established tests and hence offers promise for use as a research or clinical tool in MCI. Given the increasing numbers of smart phone, wearable, voice, and web-based computerized tests now available to assess cognition, it will also be important to directly compare these newer tests against each other at various disease stages. Such studies will allow clinicians to better personalize the tests needed for specific clinical assessments and research questions.

## Data Availability

All data produced in the present work will be made available, upon reasonable request, at the completion of our study per NIH and Columbia data sharing guidelines.

---

**Supplemental Table 1.**
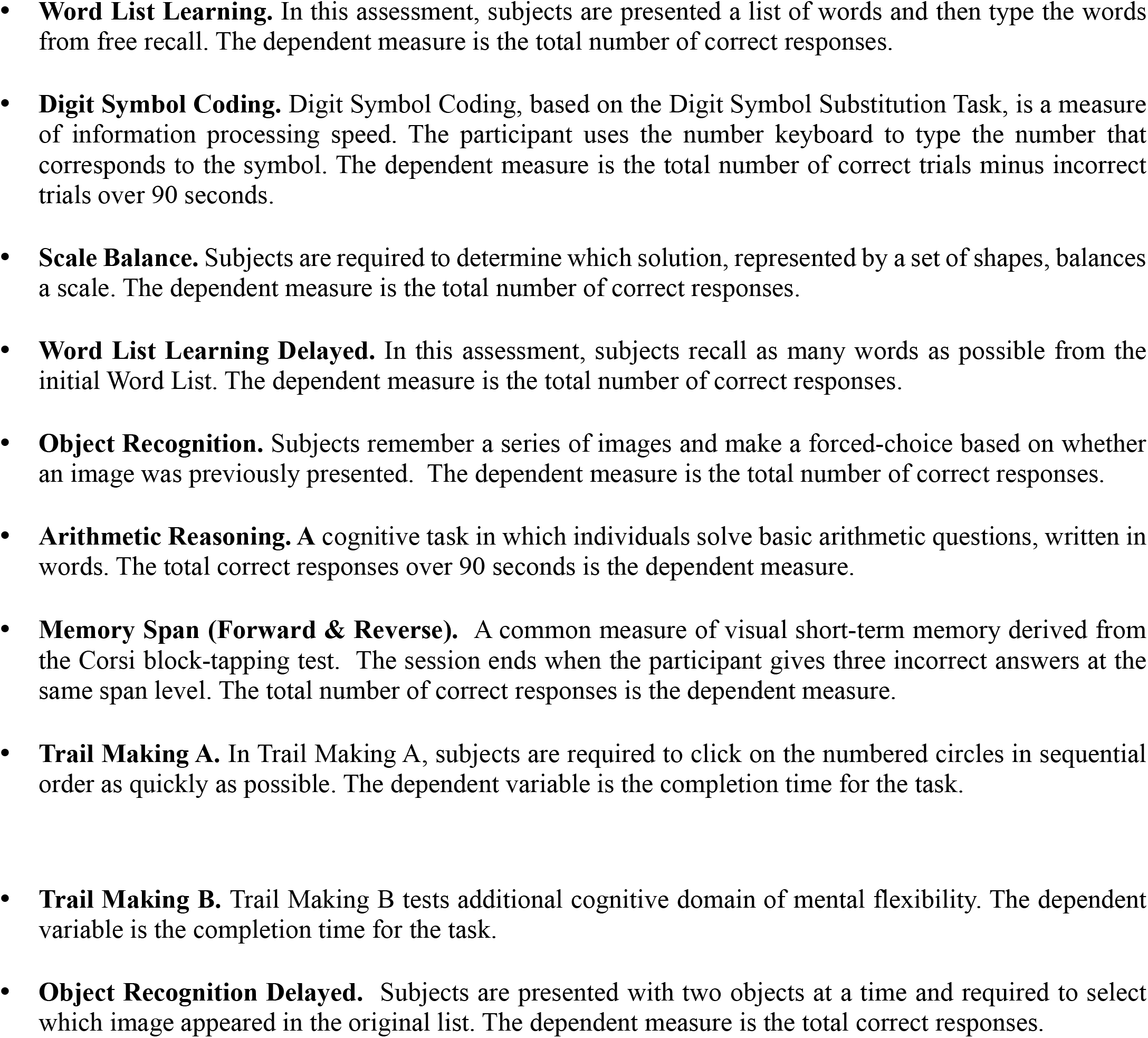
Web-Based, Self-Directed, NCPT Modules Used in This Study.

**Supplemental Table 2.** Factor Loadings of PCA with Varimax Rotation for established and NCPT measures.

|  |  | **Factor1** | **Factor2** | **Factor3** |
| --- | --- | --- | --- | --- |
| **Paper-Pencil tests** |  |  |  |  |
| ADAS-Cog |  | 0.56 | 0.43 | 0.19 |
| MMSE |  | 0.62 | 0.44 | 0.01 |
| Logical Memory immediate |  | 0.04 | 0.47 | 0.71 |
| Logical Memory delay |  | 0.22 | 0.6 | 0.55 |
| WAIS-R DSST (DSST) |  | 0.81 | 0.22 | 0.06 |
| AVLT Short Delay (AVLT-I) |  | 0.22 | 0.87 | 0.08 |
| AVLT Long Delay (AVLT-II) |  | 0.24 | 0.89 | 0.02 |
| Category Fluency |  | 0.57 | 0.16 | 0.38 |
| Boston Naming Test |  | 0.2 | -0.14 | 0.79 |
| Trail Making A |  | 0.8 | 0.11 | 0.16 |
| Trail Making B |  | 0.84 | 0.09 | 0.12 |
| **NCPT Tests** |  |  |  |  |
| Object Recognition Delay |  | 0.23 | 0.19 | 0.83 |
| Digit Symbol Coding |  | 0.61 | 0.54 | 0.27 |
| Scale Balance |  | 0.65 | 0.19 | 0.23 |
| Object Recognition Immediate |  | 0.29 | 0.21 | 0.79 |
| Memory Span |  | 0.8 | 0.03 | 0.06 |
| Trail Making A |  | 0.46 | 0.58 | -0.31 |
| Trail Making B |  | 0.72 | 0.2 | 0.23 |
| Word List Learning |  | 0.29 | 0.8 | 0.26 |
| Word List Learning Delay |  | -0.03 | 0.83 | 0.33 |
| Arithmetic Reasoning |  | 0.76 | 0.16 | 0.23 |

Table depicts factor loadings from principal component analyses with varimax rotation of 3 factors with eigen values >1. Shading indicates that factor loading was greater than 0.40. Please see text for details.

**Supplemental Table 3a:** Linear/logistic regression models based on 3 rotated factor models for NCPT measures.

|  | UPSIT (coef and p-value) |  |  | UPSA (coef and p-value) |  |  | FAQ (coef and p-value) |  |  | eMCI (odds ratio and p-value) |  |  |
| --- | --- | --- | --- | --- | --- | --- | --- | --- | --- | --- | --- | --- |
| Variable | Demo only | Factor only | full | Demo only | Factor only | full | Demo only | Factor only | full | Demo only | Factor only | full |
| Intercept | 53.42 (p=0.000) | 28.19 (p=0.000) | 41.85 (p=0.000) | 107.32 (p=0.000) | 80.90 (p=0.000) | 79.95 (p=0.000) | -1.41 (p=0.739) | 3.41 (p=0.000) | 6.62 (p=0.132) | 2.76 (p=0.647) | 0.76 (p=0.190) | 0.47 (p=0.763) |
| Factor1 |  | 0.95 (p=0.152) | 1.29 (p=0.057) |  | 4.76 (p=0.000) | 4.54 (p=0.000) |  | -1.31 (p=0.000) | -1.29 (p=0.002) |  | 1.14 (p=0.534) | 1.20 (p=0.437) |
| Factor2 |  | 3.10 (p=0.000) | 2.11 (p=0.002) |  | 4.88 (p=0.000) | 4.63 (p=0.000) |  | -1.37 (p=0.000) | -1.41 (p=0.001) |  | 1.41 (p=0.103) | 1.40 (p=0.157) |
| Factor3 |  | 2.54 (p=0.000) | 2.35 (p=0.000) |  | 3.09 (p=0.000) | 2.95 (p=0.001) |  | -0.49 (p=0.173) | -0.46 (p=0.210) |  | 1.18 (p=0.443) | 1.18 (p=0.438) |
| sex (ref. male) | 3.45 (p=0.014) |  | 3.55 (p=0.006) | 1.07 (p=0.626) |  | 1.95 (p=0.286) | -0.63 (p=0.442) |  | -0.92 (p=0.222) | 1.35 (p=0.485) |  | 1.42 (p=0.421) |
| Age | -0.32 (p=0.000) |  | -0.14 (p=0.093) | -0.50 (p=0.000) |  | -0.07 (p=0.583) | 0.12 (p=0.009) |  | -0.01 (p=0.899) | 0.98 (p=0.381) |  | 1.01 (p=0.860) |
| edu | -0.48 (p=0.026) |  | -0.56 (p=0.007) | 0.46 (p=0.179) |  | 0.15 (p=0.614) | -0.15 (p=0.226) |  | -0.08 (p=0.530) | 0.98 (p=0.786) |  | 0.97 (p=0.724) |
| R-Square | 0.286 | 0.285 | 0.439 | 0.171 | 0.44 | 0.451 | 0.087 | 0.239 | 0.252 | Pseudo R-square =0.017 | 0.037 | 0.045 |
| Adj R-Sq | 0.263 | 0.263 | 0.403 | 0.146 | 0.423 | 0.416 | 0.059 | 0.215 | 0.204 | AIC=144.59 | 142.54 | 147.66 |

**Supplemental Table 3b:** Linear/logistic regression models based on 3 rotated factor models for paper-pencil measures.

|  | UPSIT (coef and p-value) |  |  | UPSA (coef and p-value) |  |  | FAQ (coef and p-value) |  |  | eMCI (odds ratio and p-value) |  |  |
| --- | --- | --- | --- | --- | --- | --- | --- | --- | --- | --- | --- | --- |
| Variable | Demo only | Factor only | full | Demo only | Factor only | full | Demo only | Factor only | full | Demo only | Factor only | full |
| Intercept | 53.42 (p=0.000) | 28.27 (p=0.000) | 49.77 (p=0.000) | 107.32 (p=0.000) | 80.90 (p=0.000) | 98.71 (p=0.000) | -1.41 (p=0.739) | 3.41 (p=0.000) | 3.10 (p=0.394) | 2.76 (p=0.647) | 0.50 (p=0.026) | 1694.9 (p=0.051) |
| Factor1 |  | 2.13 (p=0.003) | 1.86 (p=0.007) |  | 6.69 (p=0.000) | 6.19 (p=0.000) |  | -2.05 (p=0.000) | -2.12 (p=0.000) |  | 1.29 (p=0.380) | 1.50 (p=0.272) |
| Factor2 |  | 2.33 (p=0.001) | 2.08 (p=0.002) |  | 3.71 (p=0.000) | 3.38 (p=0.000) |  | -1.31 (p=0.000) | -1.35 (p=0.000) |  | 4.19 (p=0.000) | 8.99 (p=0.000) |
| Factor3 |  | 1.23 (p=0.081) | 1.48 (p=0.017) |  | 4.22 (p=0.000) | 4.46 (p=0.000) |  | -0.65 (p=0.041) | -0.65 (p=0.047) |  | 11.11 (p=0.000) | 31.52 (p=0.000) |
| sex (ref. male) | 3.45 (p=0.014) |  | 2.86 (p=0.027) | 1.07 (p=0.626) |  | -0.37 (p=0.808) | -0.63 (p=0.442) |  | -0.24 (p=0.719) | 1.35 (p=0.485) |  | 0.83 (p=0.773) |
| Age | -0.32 (p=0.000) |  | -0.19 (p=0.022) | -0.50 (p=0.000) |  | -0.19 (p=0.051) | 0.12 (p=0.009) |  | -0.01 (p=0.831) | 0.98 (p=0.381) |  | 0.98 (p=0.652) |
| edu | -0.48 (p=0.026) |  | -0.76 (p=0.000) | 0.46 (p=0.179) |  | -0.23 (p=0.342) | -0.15 (p=0.226) |  | 0.08 (p=0.467) | 0.98 (p=0.786) |  | 0.66 (p=0.004) |
| R-Square | 0.286 | 0.195 | 0.421 | 0.171 | 0.6 | 0.623 | 0.087 | 0.396 | 0.401 | Pseudo R-square =0.017 | 0.452 | 0.513 |
| Adj R-Sq | 0.263 | 0.17 | 0.384 | 0.146 | 0.587 | 0.599 | 0.059 | 0.377 | 0.363 | AIC=144.59 | 85.55 | 79.73 |

